# Examining the Role of Maternal Religiosity in Offspring Mental Health Using Latent Class Analysis in a UK Prospective Cohort Study

**DOI:** 10.1101/2022.12.12.22283330

**Authors:** Isaac Halstead, Jon Heron, Connie Svob, Carol Joinson

## Abstract

**Background:** Previous research has examined the role of parental religious belief in offspring mental health, but has revealed inconsistent results, and suffered from a number of limitations. The aim of this study is to examine the prospective relationship between maternal religiosity and offspring mental health and psychosocial outcomes.

**Methods:** We used latent classes of religious belief (Highly religious, Moderately religious, Agnostic, Atheist) in mothers from the Avon Longitudinal Study of Parents and Children and examined their association with parent-reported mental health outcomes and self-reported psychosocial outcomes in their children at age 7-8 (n = 6079 for mental health outcomes and n = 5235 for psychosocial outcomes). We used inverse probability weighted multivariable logistic regression analysis adjusted for maternal mental health, adverse childhood experience, and socioeconomic variables.

**Results:** There was evidence for a greater risk of internalising problems among the offspring of the Highly religious and Moderately religious classes (e.g. for depression; OR = 1.51, 95% CI [1.24,1.77], OR = 1.50, 95% CI [1.26,1.73]), and greater risk of externalising problems in the offspring of the Atheist class (e.g. for ADHD; OR = 1.44, 95% CI [1.18,1.71]), compared to the offspring of the Agnostic class.

**Conclusions:** These novel findings provide evidence associations between maternal religiosity and offspring mental health differ when examined using a person-centred approach, compared to the previously used variable-centred approaches. Our findings also suggest that differences may exist in the relationship between religious (non)belief and mental health variables when comparing the UK and US.

## Introduction

Childhood mental health problems place a strain on both the child and their family (Farrell & Barrett, 2007; Houtrow & Okumura, 2011). Parental factors such as socioeconomic status, parenting style, and parental mental health are important predictors of mental health outcomes in children (Bøe et al., 2014; Leinonen et al., 2003; Manning & Gregoire, 2008; Melchior & van der Waerden, 2016). There is also some evidence for parental religiosity playing a role in offspring mental health, but the limited research that exists has found inconsistent associations (Bartkowski et al., 2008; Schottenbauer et al., 2007; Svob et al., 2018). Some studies that examined the relationship between parental religiosity and child mental health have found evidence for associations between parent religiosity and child internalising (Schottenbauer et al., 2007) and externalising problems (Bartkowski et al., 2008), and lower suicidal ideation (Svob et al., 2018). However, other studies have found a mixture of positive or no relationship (van der Jagt-Jelsma et al., 2015, 2017), or only mediated relationships (by parenting factors, with no direct association) with mental health (Kim-Spoon et al., 2012).

Some of these differences may be attributed to small sample sizes (J. Kim et al., 2009; Kim-Spoon et al., 2012; Svob et al., 2018; van der Jagt-Jelsma et al., 2017; Varon & Riley, 1999) and inadequate adjustment for confounders (Kim-Spoon et al., 2012; Schottenbauer et al., 2007; van der Jagt-Jelsma et al., 2017). These studies are also limited due to study design – of all the studies mentioned that examine parental religiosity and offspring mental health, only three use a longitudinal cohort study design, (the rest being cross-sectional) and none of these examined parental belief before the birth of the child.

Parental mental health is a potentially important confounder of the relationship between parental religiosity and child mental health. A large body of evidence indicates that parental mental health is a strong and consistent predictor of offspring mental health outcomes (Bould et al., 2015; Dean et al., 2010; M. Jacobs et al., 2012; R. H. Jacobs et al., 2015), and an individual’s religiosity is consistently, positively related to their own mental health in US samples (Braam & Koenig, 2019; Koenig, 2009). Parental socioeconomic position (SEP) is also a plausible confounder of the relationship between parental religiosity and offspring mental health. Higher parental SEP is associated with better mental and physical health in offspring (Vukojević, 2017; Cohen et al., 2009; Lemstra et al., 2008). There is also evidence for a relationship (albeit an inconsistent one) between socioeconomic variables and religiosity (Brandt & Henry, 2012; Heaton, 2013; Horowitz & Garber, 2003; Mueller & Johnson, 1975; Schieman, 2010; Schwadel, 2015; Storm, 2017; Thompson et al., 2012), which was also identified in ALSPAC (Avon Longitudinal Study of Parents and Children) (Halstead et al., 2022; Major-Smith et al., 2022). Adverse childhood experiences are also associated with religious struggles (e.g., feelings of abandonment by God) such as the death of a loved one or life-threatening events (McCormick et al., 2017), but also a desire to connect to a higher power (Santoro et al., 2016). Finally, greater parental age is simultaneously related to greater religiosity (Schwadel, 2011), and better child mental health (albeit inconsistently) (Zondervan‐Zwijnenburg et al., 2020), and should be included as a confounder.

Furthermore, previous research has been dominated by US samples. There is evidence for differences in the relationship between religious belief and mental health in the US compared to other countries, such as the UK (United Kingdom), Korea, Spain, The Netherlands, Slovenia, Estonia, Portugal, and Chile (King et al., 2013; Leurent et al., 2013; Lewis et al., 2005; Park et al., 2012). These studies have also almost exclusively used single item measures of religiosity – attendance at a place of worship, or the importance of religion in their lives. While commonly used in the religious belief literature, church attendance functions poorly when differentiating atheists and agnostics, which are both unlikely to attend church, barring weddings, funerals etc. Additionally, the importance of church attendance may also differ between religious denominations. Consequently, use of these items may be artificially constraining the variety of distinct kinds of religious (non) belief and conceal their true relationships to outcome variables.

Religiosity also relates to a variety of childhood psychosocial outcomes, such as higher self-worth (Top et al., 2003), academic achievement/scholastic competence (Jeynes, 2003; McKune & Hoffmann, 2009), and lower antisocial behaviours (Adamczyk, 2012; Laird et al., 2011; Munir & Malik, 2020). However, the role of parental religiosity has not been extensively examined (Abar et al., 2009; Bartkowski et al., 2008; Regnerus, 2003), with a mixture of positive, negative, and no associations.

The present study, based on data from a large birth cohort, the Avon Longitudinal Study of Parents and Children (ALSPAC), examines the prospective relationship between maternal religiosity and a range of child mental health outcomes (parent-reported) and psychosocial outcomes (child-reported) at age 7-8 years. This study addresses limitations of previous studies by using latent classes that describe patterns of maternal religious belief (Halstead et al., 2022) measured before the birth of the child. The latent classes describe qualitatively distinct patterns of religious belief and distinguish between highly religious, moderately religious, Agnostic and Atheist. The analysis adjusts for a range of confounders including parental mental health, parental adverse childhood events, demographic variables, and SEP indicators.

## Methods

### Participants

The Avon Longitudinal Study of Parents and Children (ALSPAC) was established to understand how genetic and environmental characteristics influence health and development in parents and children. All pregnant women resident in a defined area in the Southwest of England, with an expected date of delivery between 1st April 1991 and 31st December 1992 were invited to take part in the study. The initial number of pregnancies enrolled is 14,541. Of these initial pregnancies, there was a total of 14,676 foetuses, resulting in 14,062 live births and 13,988 children who were alive at 1 year of age. These parents and children have been followed over the last 30 years and have completed a variety of questionnaires concerning their demographics, physiological and genetic data, life events, physical, and psychological characteristics. For more information, see Boyd et al., 2013; Fraser et al., 2013. Please note that the study website contains details of all the data that is available through a fully searchable data dictionary and variable search tool (http://www.bristol.ac.uk/alspac/researchers/our-data/).

**Figure 1.**
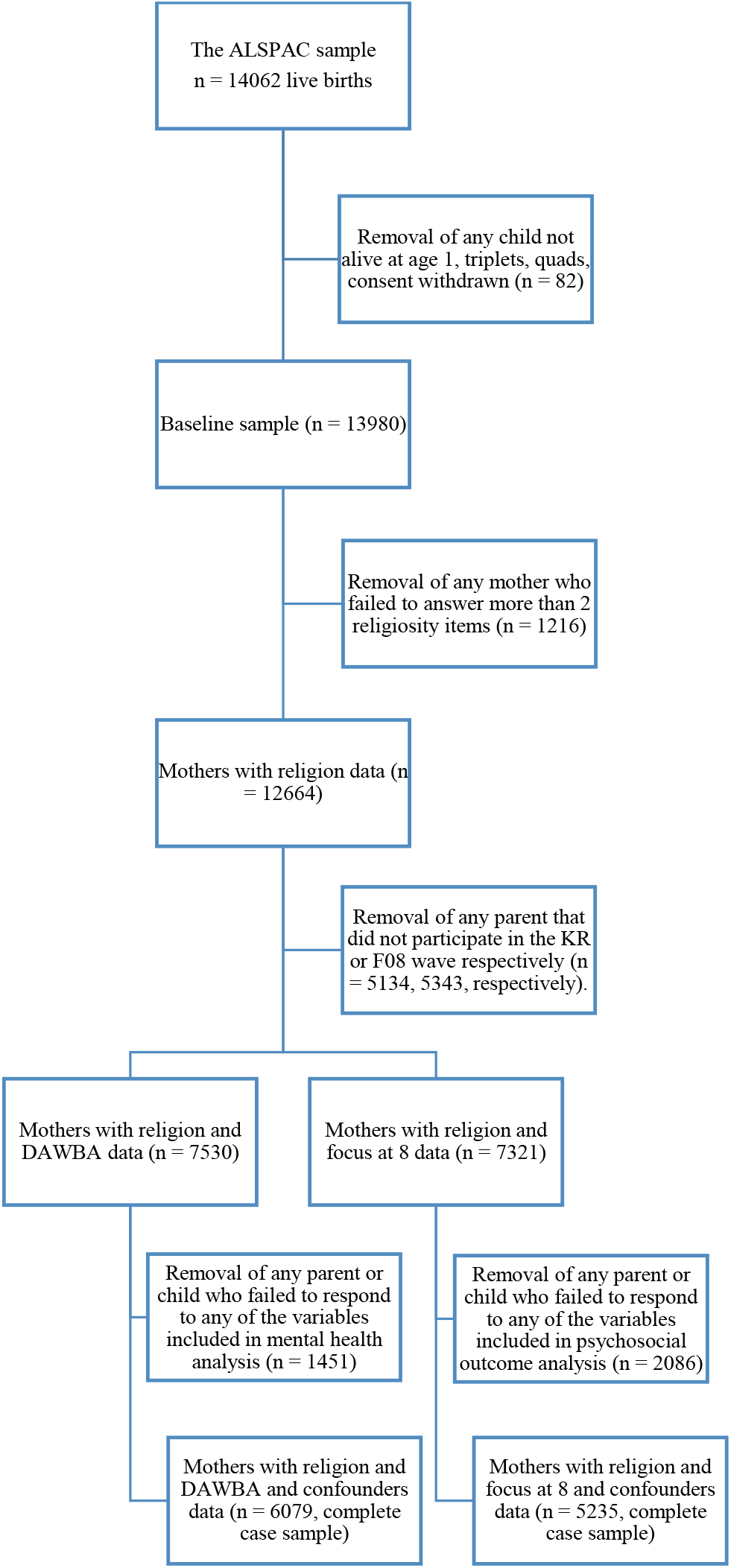
Sample flowchart showing each stage of exclusion.

## Exposures

### Latent classes of maternal religiosity

We used the maternal latent class membership variables derived by Halstead et al., (2022) as our indicators of religiosity in the mothers of the ALSPAC parent cohort, based upon the assumptions that they would be the primary caregiver, the classes are stable over time (D. Smith et al., 2022) and that maternal and paternal religious latent classes are associated (see Halstead et al., 2022). The latent classes are composed of a series of conditional probabilities, which are used to label and describe the classes. These classes provide a more nuanced alternative to variable centred approaches that measure religiosity using a single item by providing qualitatively distinct types of religious belief rather than a simple continuum. The questions used to derive the classes include belief in God, whether a person has asked for help from God, whether they would ask for God’s help when in trouble, the duration of their faith, their church attendance, and whether they have received help from individuals from their own or another religion, which were measured at the antenatal timepoint. Our choice to use the latent classes generated at the antenatal timepoint were motivated by the larger sample size provided by this timepoint. This decision is also supported by the small amount of transition between classes over time (see D. Smith et al., 2022 for a descriptive account of these transitions). The classes were named the Highly Religious, Moderately Religious, Agnostic, and Atheist and each represented approximately 14%, 30%, 38%, and 18% of the sample, respectively. For details of these questions, see Table 2 of the appendix.

### Outcomes

#### Parent-reported child mental health outcomes

When their study child was aged 7 years, mothers were asked to complete the Development and Wellbeing Assessment (DAWBA) (Goodman et al., 2000) which includes questions about symptoms of common mental health disorders. We included symptoms of separation anxiety, phobias, social anxiety, obsessive compulsive disorder (OCD), generalised anxiety disorder (GAD), depression, attention deficit hyperactivity disorder (ADHD), oppositional defiant disorder (ODD) and conduct disorder. A small number of children met DSM-IV criteria for psychiatric disorders in the ALSPAC cohort at this age (Joinson et al., 2006). We therefore created binary variables to indicate the presence of any symptom that was severe, e.g., rated as “a lot more than others” or “a great deal” (See Table 1 of the appendix for details of items used). Only the complete case samples were used for analyses. The prevalence of mental health symptoms is provided in Table S3 of the appendix.

**Table 1.** Distribution of confounders across the samples

| Sample size |  | Baseline (up to<br>13980) | Mothers with<br>religion data<br>(12664) | Religion and<br>DAWBA (7530) | Religion and<br>DAWBA and<br>confounders (6079) | Religion and focus<br>at 8 (7321) | Religion and Focus<br>at 8 and<br>confounders (5235) |
| --- | --- | --- | --- | --- | --- | --- | --- |
| Social class (Ref;<br>non-manual) | Manual | 2933 (25.6%) | 2611 (24.6%) | 1401(21.1%) | 1235 (20.3%) | 1353 (21.0%) | 1038 (19.8%) |
| Financial problems<br>(Ref; no) | Yes | 1510 (12.1%) | 1319 (11.6%) | 665(9.6%) | 569 (9.4%) | 673 (10.0%) | 510 (9.7%) |
| Financial hardship<br>(Ref; no) | Yes | 3298 (26.1%) | 2970 (25.3%) | 1590 (22.0%) | 1265 (20.8%) | 1494 (21.3%) | 1026 (19.9%) |
| Education (Ref; A-level or higher) | None/vocational | 3869 (29.8%) | 3381 (28.2%) | 1621 (21.9%) | 1259 (20.7%) | 1524 (21.3%) | 1019 (19.5%) |
|  | O Level | 4485 (34.6%) | 4243 (35.4%) | 2648 (35.8%) | 2228 (36.7%) | 2528 (35.3%) | 1875 (35.8%) |
| Home status (Ref;<br>bought) | Rented | 3578 (26.4%) | 2941 (24.0%) | 1282 (17.4%) | 893 (14.7%) | 1136 (15.8%) | 695 (13.3%) |
| ACE (Ref; 0 ACE) | 1 | 2026 (20.4%) | 2330 (18.4%) | 1479 (19.6%) | 1174 (19.3%) | 1424 (19.5%) | 988 (18.7%) |
|  | 2 | 1387 (14.0%) | 1419 (11.2%) | 860 (11.4%) | 684 (11.3%) | 813 (11.1%) | 573 (10.8%) |
| Maternal age at<br>baseline | Mean (SD) | 28.4 (4.8) | 28.5 (4.7) | 29.1 (4.5) | 29.3 (4.4) | 29.3 (4.5) | 29.5 (4.4) |
| Maternal<br>depression | Median (IQR) | 7 (4.9) | 6.9 (4.8) | 6.5 (4.6) | 6 (6) | 6.5 (4.6) | 6 (6) |
| Maternal anxiety | Median (IQR) | 4.9 (3.5) | 4.9 (3.5) | 4.7 (3.4) | 4 (4) | 4.7 (3.4) | 4 (4) |

Note. The samples in the table correspond to the samples in the flowchart.

### Self-reported psychosocial outcomes

Child self-reported psychosocial outcomes were obtained from a ‘Focus Clinic’ attended by children when they were aged 8 years. The prevalence of psychosocial outcomes is provided Table S4 of the appendix. All reported Cronbach’s α refer to the current paper, using the complete case samples.

#### Bullying (as the bully and victim)

We assessed peer victimisation through self-report using a modified version of the bullying and friendship interview schedule (Wolke et al., 2001). The scale consists of measures of being an overt victim (α = 0.593), relational victim (α = 0.623), overt bully (α = 0.510), and relational bully (α = 0.447).

#### Scholastic competence and global self-worth

This measure used a 12-item version of Harter’s Self Perception Profile for Children (SPPC) (Harter, 1985) comprising global self-worth (α = 0. 651) and scholastic competence (α = 0. 691), rather than the full 36 item scale that contains the other subscales.

#### Unhappy with friends

A series of five questions taken from the Cambridge Hormones and Moods Project Friendship Questionnaire (Goodyer et al., 1990) to indicate unhappiness with friends (α = 0. 503).

#### Antisocial activities

This measure used 15 questions, including 11 from the Self-Reported Antisocial Behaviour for Young Children Questionnaire (Loeber et al., 1989), three dummy questions and an additional example question, to measure antisocial activities (α = 0. 582).

### Confounders

We chose confounders based upon empirical evidence of a relationship with the exposure and outcome variables. Confounders were assessed by maternal reports in the antenatal period and included maternal age at baseline, maternal mental health, and SEP indicators. We also adjusted for retrospective reports of maternal adverse childhood experiences (ACEs) assessed in questionnaires completed during the antenatal period and when the study child was aged 2 years. Details of the confounding variables are provided in Table S1 of the appendix.

#### Statistical analyses

We used logistic regression to calculate odds ratios and 95% confidence intervals for each mental health and psychosocial variable and their association with maternal religious latent class, both before and after adjustment for confounders. As the outcomes we are examining are rare (⪅10%) we interpreted the effect estimates as risk ratios, and we attributed changes seen to parameter estimates in the multivariable models as being due to confounding (Zammit et al., 2002). Risk ratios were estimated in relation to the Agnostic class, as this was the largest class in the sample, and was characterised by the most ‘moderate’ beliefs (i.e. neither religious nor atheist). Parameter estimates were then adjusted for confounders. The dataset was constructed in R studio (R Core Team, 2021) and all analyses were carried out in Mplus (Version 8.7), using a bias adjusted 3 step latent class analysis which incorporates uncertainty in latent class assignment (Vermunt & Magidson, 2021).

#### Weighted estimates

To address potential bias due to missing data, we created weighting variables based on SEP variables as SEP is associated with attrition in the ALSPAC sample (Fernández-Sanlés et al., 2021; Howe et al., 2013). This was done by using inverse probability weighting, using SEP indicators with minimal missingness (less than 5%) including home ownership status, cigarette smoking, car ownership, and education, with any missingness recoded in these variables to be the modal response category. These were then used as exposures in a logistic regression model, with missingness at the 7-year and 8-year timepoints (when our outcome variables were measured) as the outcome variables. From these models, the weights were calculated and added to the main dataset, for use in the weighted analyses. The results of the weighted analyses are presented as the main results and the unweighted results are provided in Table S7 and S8 of the appendix. Additionally, full details of each adjusted model may be found in Table S5 and S6 of the appendix.

## Results

Compared with the baseline sample, the restricted sample used in the complete case analysis comprised a higher proportion of participants of higher SEP (i.e. homeowners, non-manual social class, no major financial problems, and no financial hardship). The restricted sample also had a lower proportion of maternal depression, anxiety and adverse childhood events. See Table 1 for more details.

### Association between maternal religiosity latent classes and parent-reported child mental health outcomes

Table 2 shows the results of the logistic regressions with parent-reported mental health variables as outcomes. In the unadjusted models, children of mothers in the **Highly Religious** class, compared with the Agnostic class have increased risk of ADHD, depression, OCD, and ODD. Compared with the Agnostic class, children of mothers in the **Moderately Religious** class have increased risk of depression, anxiety, and ODD. Compared with the Agnostic class, children of mothers in the **Atheist** class have increased risk of ADHD, conduct disorder, and ODD. The highest level of attenuation in the adjusted models was found for conduct disorder, with a 9% reduction in the risk ratio in the fully adjusted model. There was little evidence of confounding in the adjusted models for the other mental health symptoms.

**Table 2.** Weighted risk ratios and 95% confidence intervals for the associations between maternal religious latent class and offspring mental health at age 7, with the Agnostic class as the reference class.

|  | OR (CI)<br>Model 1 | OR (CI)<br>Model 5 |  | OR (CI)<br>Model 1 | OR (CI)<br>Model 5 |
| --- | --- | --- | --- | --- | --- |
| **ADHD** |  |  | **Social phobia** |  |  |
| Agnostic | 1.00 ref | 1.00 ref | Agnostic | 1.00 ref | 1.00 ref |
| Highly religious | 1.29(1.00,1.58) | 1.34(1.05,1.64) | Highly religious | 1.39(1.01,1.76) | 1.33(0.95,1.71) |
| Moderately religious | 1.16(0.90,1.41) | 1.17(0.91,1.43) | Moderately religious | 1.25(0.91,1.58) | 1.24(0.90,1.57) |
| Atheist | 1.44(1.18,1.71) | 1.41(1.14,1.68) | Atheist | 1.04(0.65,1.43) | 1.01(0.62,1.40) |
| P - value | .043 | .053 | P - value | .255 | .328 |
| **Conduct disorder** |  |  | **Specific phobia** |  |  |
| Agnostic | 1.00 ref | 1.00 ref | Agnostic | 1.00 ref | 1.00 ref |
| Highly religious | 1.35(0.98,1.72) | 1.37(0.99,1.75) | Highly religious | 1.01(0.74,1.28) | 1.06(0.78,1.33) |
| Moderately religious | 1.19(0.86,1.53) | 1.22(0.89,1.56) | Moderately religious | 1.03(0.80,1.26) | 1.05(0.82,1.28) |
| Atheist | 1.55(1.22,1.89) | 1.46(1.12,1.80) | Atheist | 0.86(0.60,1.12) | 0.84(0.58,1.11) |
| P - value | .067 | .133 | P - value | .585 | .383 |
| **Depression** |  |  | **ODD** |  |  |
| Agnostic | 1.00 ref | 1.00 ref | Agnostic | 1.00 ref | 1.00 ref |
| Highly religious | 1.51(1.24,1.77) | 1.40(1.13,1.68) | Highly religious | 1.73(1.37,2.10) | 1.72(1.35,2.08) |
| Moderately religious | 1.50(1.26,1.73) | 1.48(1.24,1.71) | Moderately religious | 1.39(1.06,1.72) | 1.38(1.04,1.72) |
| Atheist | 1.17(0.90,1.44) | 1.13(0.86,1.40) | Atheist | 1.44(1.08,1.79) | 1.39(1.02,1.75) |
| P - value | .001 | .004 | P - value | .020 | .027 |
| **General anxiety** |  |  | **Separation anxiety** |  |  |
| Agnostic | 1.00 ref | 1.00 ref | Agnostic | 1.00 ref | 1.00 ref |
| Highly religious | 1.21(0.89,1.53) | 1.22(0.90,1.55) | Highly religious | 1.19(0.83,1.55) | 1.18(0.81,1.56) |
| Moderately religious | 1.44(1.18,1.70) | 1.43(1.17,1.70) | Moderately religious | 1.23(0.92,1.54) | 1.25(0.94,1.56) |
| Atheist | 1.24(0.94,1.53) | 1.24(0.94,1.53) | Atheist | 1.09(0.75,1.44) | 1.05(0.70,1.40) |
| P - value | .056 | .066 | P - value | .552 | .775 |
| **OCD** |  |  |  |  |  |
| Agnostic | 1.00 ref | 1.00 ref |  |  |  |
| Highly religious | 1.60(1.21,2.00) | 1.53(1.13,1.92) |  |  |  |
| Moderately religious | 0.65(0.19,1.11) | 0.64(0.18,1.10) |  |  |  |
| Atheist | 1.04(0.63,1.46) | 1.03(0.62,1.45) |  |  |  |
| P - value | .004 | .008 |  |  |  |

**Note.** *Model 1* is the unadjusted model, *Model 5* adjusts for maternal age, SEP, ACE, and maternal mental health. P values are omnibus p-values based on a Wald test with 3 d.f.

### Association between maternal religiosity latent classes and self-reported psychosocial outcomes

Table 3 that compared with the Agnostic class, children of mothers in the **Highly Religious** class have increased risk of antisocial behaviour, and decreased risk of being an overt bully. Compared with the Agnostic class, children of mothers in the **Moderately Religious** class have increased risk of self-reported antisocial behaviour and low scholastic competence. Compared with the Agnostic class, children of mothers in the **Atheist** class have increased risk of self-reported antisocial behaviour, but lower risk of being overt bullies. The highest level of attenuation in the adjusted models was found for low scholastic competence, with an 8% reduction in the risk ratio in the fully adjusted model. There was little evidence of confounding in the adjusted models for the other psychosocial outcomes.

**Table 3.** Weighted risk ratios and 95% confidence intervals for the associations between maternal religious latent class and offspring psychosocial outcomes at age 8, with the Agnostic class as the reference class.

|  | OR (CI)<br>Model 1 | OR (CI)<br>Model 5 |  | OR (CI)<br>Model 1 | OR (CI)<br>Model 5 |
| --- | --- | --- | --- | --- | --- |
| **Antisocial behaviour** |  |  | **Overt bully** |  |  |
| Agnostic | 1.00 ref | 1.00 ref | Agnostic | 1.00 ref | 1.00 ref |
| Highly religious | 1.29(1.06,1.52) | 1.29(1.06,1.52) | Highly religious | 0.60(0.22,0.98) | 0.59(0.21,0.98) |
| Moderately religious | 1.24(1.04,1.44) | 1.24(1.04,1.44) | Moderately religious | 0.74(0.38,1.09) | 0.73(0.38,1.09) |
| Atheist | 1.42(1.21,1.64) | 1.40(1.18,1.61) | Atheist | 0.57(0.20,0.94) | 0.58(0.21,0.95) |
| P - value | .008 | .013 | P - value | .013 | .013 |
| **Low scholastic competence** |  |  | **Relational bully** |  |  |
| Agnostic | 1.00 ref | 1.00 ref | Agnostic | 1.00 ref | 1.00 ref |
| Highly religious | 0.96(0.74,1.17) | 1.04(0.82,1.26) | Highly religious | 0.71(0.13,1.29) | 0.65(0.07,1.24) |
| Moderately religious | 1.17(1.00,1.35) | 1.22(1.04,1.40) | Moderately religious | 1.03(0.48,1.59) | 1.05(0.48,1.61) |
| Atheist | 1.10(0.90,1.30) | 1.12(0.92,1.32) | Atheist | 0.90(0.30,1.49) | 0.90(0.31,1.49) |
| P - value | .241 | .184 | P - value | .642 | .458 |
| **Low self-worth** |  |  | **Relational victim** |  |  |
| Agnostic | 1.00 ref | 1.00 ref | Agnostic | 1.00 ref | 1.00 ref |
| Highly religious | 1.26(0.99,1.52) | 1.27(1.00,1.54) | Highly religious | 0.77(0.52,1.02) | 0.74(0.49,0.99) |
| Moderately religious | 1.15(0.91,1.38) | 1.17(0.94,1.41) | Moderately religious | 0.94(0.72,1.16) | 0.92(0.70,1.15) |
| Atheist | 1.17(0.91,1.43) | 1.16(0.90,1.42) | Atheist | 1.08(0.82,1.33) | 1.09(0.84,1.35) |
| P - value | .324 | .289 | P - value | .103 | .045 |
| **Overt victim** |  |  | **Unhappy with friends** |  |  |
| Agnostic | 1.00 ref | 1.00 ref | Agnostic | 1.00 ref | 1.00 ref |
| Highly religious | 0.91(0.71,1.11) | 0.89(0.69,1.10) | Highly religious | 1.15(0.91,1.39) | 1.15(0.91,1.40) |
| Moderately religious | 0.95(0.78,1.12) | 0.95(0.78,1.12) | Moderately religious | 0.93(0.72,1.14) | 0.94(0.72,1.15) |
| Atheist | 0.94(0.75,1.14) | 0.96(0.76,1.15) | Atheist | 0.77(0.51,1.02) | 0.76(0.51,1.01) |
| P - value | .827 | .743 | P - value | .047 | .043 |

**Note.** *Model 1* is the unadjusted model, *Model 5* adjusts for maternal age, SEP, ACE, and maternal mental health. P values are omnibus p-values based on a Wald test with 3 d.f.

## Discussion

The current study found evidence that maternal religious belief was associated with a range of mental health and psychosocial outcomes in their offspring at age 7-8 years. Compared with children of Agnostic mothers, the children of Highly religious and Moderately religious parents were at greater risk of internalising symptoms, and the children of Atheist parents were at greater risk of externalising symptoms. However, there was no clear pattern of results for psychosocial outcomes. These associations were independent of maternal age, SEP variables, ACE, and mental health. There were only a few instances of attenuation, with most of the results being robust to the inclusion of confounding variables.

### Strengths and limitations

There are several strengths to this study. The use of data from a large prospective community based cohort, use of latent classes of belief, rather than relying upon single item measures which dominate the religiosity literature, availability of parent- and child-reported mental health and psychosocial outcomes based on validated questionnaires, and availability of data on a wide range of important confounders The use of latent classes provided insights that may have otherwise been lost by using items that fail to differentiate between Agnostic and Atheist individuals, such as through the use of items that ask participants to indicate the importance of religion in their lives or church attendance.

There are also limitations that should be considered when interpreting the findings. There was a large amount of attrition between the baseline sample, and the final complete case analysis which could lead to selection bias. Specifically, those with higher SEP (Howe et al., 2013) and religiosity (Morgan et al., 2022) are more likely to participate in ALSPAC initially and continue to participate over time. Attrition is strongly related to low SEP in ALSPAC, and the present study attempted to mitigate this using weighting to account for the potential bias, in line with recommendations (Howe et al., 2013).

### Comparisons with previous research

Our findings are contrary to previous research which examined the relationship between parental religious belief and child mental health, which has found that greater parental (either maternal or paternal) religious belief is associated with better mental health outcomes in offspring (Svob et al., 2018; Varon & Riley, 1999). A possible reason for this difference is that the current study is based on a UK sample, compared to previous research which has predominantly been based on US samples (e.g., 75% of the religious belief and mental health research reviewed by Koenig, 2009 was conducted on US samples). There is evidence the UK differs in its relationship with religious belief and mental health compared to other countries, with some studies finding that increased religious or spiritual belief was associated with worse mental health outcomes such as depression, anxiety, and phobias (King et al., 2013; Leurent et al., 2013). Additionally, given that the US is highly religious compared to the UK, and most previous studies contain relatively small numbers of individuals that could be considered atheist or agnostic, previous studies may be less suited to capturing the differences between them and religious individuals.

The use of items that assume religious belief exists on a simple continuum from non-religious to highly religious may be obscuring the true nature of religion’s relationship with mental health and psychosocial outcomes. The relationship between religious belief and mental health may be better explained by using qualitatively different types of belief in future research. Given previous literature’s inclination to use items that ask about religious attendance or the importance of religion, and both Atheists and Agnostics are likely to respond the same way to these items (i.e. they are both unlikely to attend church), previous research may be failing to acknowledge the way these items function for different groups. Conflating Atheist and Agnostic groups appears to hide important differences in outcomes between the two.

### Possible mechanisms explaining the current findings

The increased mother-reported externalising problems and psychosocial outcomes in the Highly Religious mothers (e.g. ADHD, ODD, antisocial behaviour) may be partially explained by highly religious mothers having higher expectations of their child’s morals (Rhodes & Nam, 1970; C. Smith, 2003), an increased likelihood to perform parental monitoring activities (Guo, 2018; Y.-I. Kim & Wilcox, 2014) and to be more engaged in their child’s life (Guo, 2018). This possibly leads them to be more attentive to ADHD or ODD symptoms, as well as negative (e.g. antisocial) behaviours. However, children of religious parents also perceive their parents to be more controlling, which could in turn lead to more internalising and externalising problems (Bornstein et al., 2017). Combined, this suggests that a degree of monitoring or control is healthy and may lead to more attentive parenting, but when it is perceived to be excessive, it can be a stressor to the child. In the context of the current study, the higher degree of control that religious mothers exert over their children may impact their levels of stress, leading to a greater risk of depression or anxiety (Bean & Catania, 2018; Rapee, 1997; Yap et al., 2014). The increased mother reported internalising symptoms may be explained by children of religious parents being religious themselves, which may incline them towards rumination, which leads to greater internalising symptoms (Saunders et al., 2021). Religious belief is associated with OCD (Abramowitz et al., 2004; Himle et al., 2011; Sica et al., 2002), possibly through heightened pathogen disgust sensitivity (Olatunji et al., 2007), which is also associated with religiosity (Stewart et al., 2020; Yu et al., 2021). Consequently, the behaviours associated with OCD symptoms may be taught to offspring during their upbringing (Waters & Barrett, 2000). There is little existing literature to explain the pattern of results in the Atheist class, or for the differences between the Moderately and Highly Religious classes. We may nevertheless speculate that the association between spirituality, depression, and other internalizing disorders may be partially explained by an increased interior-orientation, observed for example in rumination (Saunders et al., 2021) and default-mode network connectivity (Svob et al., 2016) in those who are spiritual. Whereas, Atheism may support a more external worldview and contribute to its greater likelihood to be associated with externalizing disorders. Further research is needed to examine the replicability of our findings in non-US samples.

### Conclusions

Contrary to existing literature, we found evidence that maternal religiosity is associated with a higher risk of internalising symptoms. Children of atheist parents are at greater risk of externalising symptoms. Future research is needed to determine whether these relationships are causal and to identify the underlying mechanisms. For example, it is conceivable that the relationship may be mediated by parenting style/quality variables or moderated by partner religious class.

## Data Availability

Data Availability
Please see the ALSPAC data management plan which describes the policy regarding data sharing (http://www.bristol.ac.uk/alspac/researchers/data-access/documents/alspac-data-management-plan.pdf), which is by a system of managed open access. Data used for this submission will be made available on request to the Executive. The datasets presented in this article are linked to ALSPAC project number B4001, please quote this project number during your application.
The steps below highlight how to apply for access to the data included in this study and all other ALSPAC data:
1. Please read the ALSPAC access policy (http://www.bristol.ac.uk/media-library/sites/alspac/documents/researchers/data-access/ALSPAC_Access_Policy.pdf) which describes the process of accessing the data and samples in detail, and outlines the costs associated with doing so.
2. You may also find it useful to browse our fully searchable research proposals database (https://proposals.epi.bristol.ac.uk/?q=proposalSummaries), which lists all research projects that have been approved since April 2011.

http://www.bristol.ac.uk/media-library/sites/alspac/documents/researchers/data-access/ALSPAC_Access_Policy.pdf

## Ethical standards

Ethical approval for the study was obtained from the ALSPAC Ethics and Law Committee and the Local Research Ethics Committees. Informed consent for the use of data collected via questionnaires and clinics was obtained from participants following the recommendations of the ALSPAC Ethics and Law Committee at the time.

## Acknowledgements

We are extremely grateful to all the families who took part in this study, the midwives for their help in recruiting them, and the whole ALSPAC team, which includes interviewers, computer and laboratory technicians, clerical workers, research scientists, volunteers, managers, receptionists, and nurses.

## Data Availability

Please see the ALSPAC data management plan which describes the policy regarding data sharing (http://www.bristol.ac.uk/alspac/researchers/data-access/documents/alspac-data-management-plan.pdf), which is by a system of managed open access. Data used for this submission will be made available on request to the Executive. The datasets presented in this article are linked to ALSPAC project number B4001, please quote this project number during your application.

The steps below highlight how to apply for access to the data included in this study and all other ALSPAC data:

1. Please read the ALSPAC access policy (http://www.bristol.ac.uk/media-library/sites/alspac/documents/researchers/data-access/ALSPAC_Access_Policy.pdf) which describes the process of accessing the data and samples in detail, and outlines the costs associated with doing so.
2. You may also find it useful to browse our fully searchable research proposals database (https://proposals.epi.bristol.ac.uk/?q=proposalSummaries), which lists all research projects that have been approved since April 2011.
3. Please submit your research proposal (https://proposals.epi.bristol.ac.uk/) for consideration by the ALSPAC Executive Committee. You will receive a response within 10 working days to advise you whether your proposal has been approved.

