## Supplementary material for "Examining the Role of Maternal Religiosity in Offspring Mental Health Using Latent Class Analysis in a UK Prospective Cohort Study": Mental health LCA appendix 05-12-22.pdf

**Table S1.** Measures used for analysis.

| **Measure/questionnaire** | **Item wording** | **Response options** | **Reporter and format of reporting** | **Derivation of measure used in analysis** | **Variable name** |
|---|---|---|---|---|---|
| **Child variables** | | | | | |
| Childhood mental health (DAWBA) | Please compare her behaviour in the last 6 months with other children of her age.<br><br>Does she often fidget?<br><br>Is it hard for her to stay sitting down for long?<br><br>Does she run or climb about when she shouldn't?<br><br>Does she find it hard to play or take part in other leisure activities without making a noise?<br><br>If she is rushing about does she find it hard to calm down when someone asks her to do so ? | 1; No more than others<br>2; A little more than others<br><br>3; A lot more than others | Mother, questionnaire | Only the children who were reported to have suffered from the symptoms the most often, or a lot more than others of the same age were included in the group with internalizing or externalizing problems (with the exception of 'any' mood symptoms because the options in the symptom scale were 'yes/no'). | kr201<br>kr202<br>kr203<br>kr204<br>kr205<br>kr206<br>kr207<br>kr208<br>kr209<br>kr210<br>kr251<br>kr252<br>kr253<br>kr254<br>kr255<br>kr256<br>kr225<br>kr226<br>kr227<br>kr228<br>kr229<br>kr230<br>kr231<br>kr357<br>kr358<br>kr359<br>kr360<br>kr361<br>kr362<br>kr363<br>kr410<br>kr411<br>kr412<br>kr413<br>kr414<br>kr415<br>kr416 |

| | | | | | |
|---|---|---|---|---|---|
| | | | | | kr417<br>kr418<br>kr419<br>kr420<br>kr436<br>kr437<br>kr438<br>kr439<br>kr440<br>kr441<br>kr442<br>kr443<br>kr444<br>kr481<br>kr482<br>kr483<br>kr484<br>kr485<br>kr486<br>kr487<br>kr488<br>kr489<br>kr503<br>kr505<br>kr507<br>kr509<br>kr511<br>kr513<br>kr515 |
| Bullying | ***Overt bully/victim***<br>Has child hit/beaten up others/been hit/beaten up?<br><br>***Relational bully/victim***<br>Has child spoilt other childrens' games/had games spoilt? | 1; 1-3 times in the past 6 mths<br>2; 4+ times last 6 mths but <1/wk<br>3; At least 1/wk<br>4; Child refused<br>5; Ch said DK | Child; questionnaire | If child responded *yes* to any bullying event he/she will be classified as overt or relational bully/victim if bullying event happened frequently (several times in 6 months) or very frequently (several times a week). | Overt victim<br>F8FP140<br>F8FP150<br>F8FP160<br>F8FP170<br>F8FP180<br>Relational victim<br>F8FP330<br>F8FP340<br>F8FP350<br>F8FP360<br>Overt bully<br>F8FP240<br>F8FP250<br>F8FP260<br>F8FP270 |

| | | | | | |
|---|---|---|---|---|---|
| | | | | | F8FP280<br>Relational bully<br>F8FP410<br>F8FP420<br>F8FP430 |
| Self-worth | Some children are happy with themselves as a person. | 5;Yes, really like me;<br>4;Sort of true for me;<br>3;Yes, a bit like me;<br>2;No, not really like me;<br>1;No, not at all like me. | Child; questionnaire | This is a derived variable from ALSPAC.<br>Scores ranged from 6–24; scores equal to or less than 14 will be coded as low scores. | f8se111<br>f8se113<br>f8se115<br>f8se117<br>f8se119<br>f8se121 |
| Scholastic competence | Some children do very well at their classwork. | 5;Yes, really like me;<br>4;Sort of true for me;<br>3;Yes, a bit like me;<br>2;No, not really like me;<br>1;No, not at all like me. | Child; questionnaire | This is a derived variable from ALSPAC.<br>Scores ranged from 6–24; scores equal to or less than 14 will be coded as low scores. | f8se110<br>f8se112<br>f8se114<br>f8se116<br>f8se118<br>f8se120 |
| Unhappy with friends | Are you happy with the number of friends you've got? | 1, Very happy,<br>2, Quite happy,<br>3, Quite unhappy,<br>4, Unhappy,<br><br>And<br>1, Almost every day,<br>2, More than once week,<br>3, Less than once week,<br>4, Hardly ever,<br><br>With an option to say 'dont know' | Child; questionnaire | This is a derived variable from ALSPAC.<br>Scores ranged from 0–15; with a higher score indicating less happiness with friendship. A score of 6 or more was coded as a high score. | f8fs110<br>f8fs111<br>f8fs112<br>f8fs113<br>f8fs114 |
| Antisocial activities | Examples include Have you ever taken something from a shop without paying for it? and Have you ever tried a cigarette? | 1 = never 2= ever | Child; questionnaire | This is a derived variable that is based upon the total number of activities the child reported doing from 11 different items. | f8aa101<br>f8aa102<br>f8aa103<br>f8aa105<br>f8aa107<br>f8aa108<br>f8aa110<br>f8aa111<br>f8aa112<br>f8aa113<br>f8aa114 |

| **Measure** | **Item wording** | **Response options** | **Reporter and format of reporting** | **Derivation of measure used in analysis** | **Variable name** |
|---|---|---|---|---|---|
| **Parent variables** | | | | | |
| Anxiety | Do you feel upset for no obvious reason? | 1; Often to 4; Never. | Mother; questionnaire | The 18 weeks scores will be used for analysis. | b351 |
| Depression | I have looked forward with enjoyment to things | 1; As much as I ever did to 4; Hardly at all. | Mother; questionnaire | The 18 weeks scores will be used for analysis. | b370 |
| Social class | | 1; Professional<br>2; Managerial and technical<br>3; Skilled non-manual<br>4; Skilled manual<br>5; Partly skilled<br>6; Unskilled | Mother; questionnaire | This scale was recoded into a binary variable of either manual (partly, or unskilled occupations) or non-manual work (professional, managerial, or skilled professions) | c_sc_m |
| Education | | 5 categories:<br>1; vocational,<br>2; certificate of secondary education (CSE),<br>3;O-level,<br>4; A-level,<br>5; Degree. | Mother; questionnaire | This variable was recoded as 1; CSE *or vocational*, 2; *A-level or O-level*, 3; *Degree*. | c645a |
| Financial problems | | 1; affected a lot<br>2; fairly affected<br>3; mildly affected<br>4; No effect at all<br>5; didnt happen | Mother; questionnaire | This variable was recoded into a binary, to indicate either affected by financial problems or not at all/didnt happen | b594 |
| Financial hardship | Measured using a variable derived from items that asked participants whether they had difficulty in affording food, clothing, heating, rent or mortgage, and things they will need for their baby | This derived variable is measured on a 15-point scale, with a higher score indicating a higher level of financial hardship. | Mother; questionnaire | This variable will be recoded into a binary measure using a cut-off of ≥5 which corresponds to ≈20% of the highest financial hardship scores in the sample. | c525 |

| | | | | | |
|---|---|---|---|---|---|
| Housing status | Is your home: | 1; Being bought/mortgaged<br>2; being bought from council<br>3; owned - with no mortgage to pay<br>4; rented from council<br>5; rented from private<br>6; landlord – furnished<br>7; rented from private landlord – unfurnished<br>8; rented from housing association<br>9; other (please describe) | Mother; questionnaire | This will be recoded into a binary variable of either rented or bought accommodation | a006 |
| Age | | | Mother; questionnaire | | a901 |
| Parental adverse childhood experiences | Sexual abuse<br>Non-sexual abuse<br>Maladaptive family functioning<br>Parental mental illness<br>Maternal lack of care<br>Maternal overprotection | | Mother; questionnaire | ACE scores derived using an approach adapted from (Crick et al., 2022) | c850<br>c870<br>c910<br>c925<br>c950<br>c420<br>c416<br>h134<br>h135<br>d755<br>c421a<br>h137a<br>h137c<br>h137d<br>h137b<br>h137e<br>h137f<br>h137g<br>h137h<br>d387<br>c423<br>d537<br>d536<br>d529<br>d528<br>d578<br>d579<br>d701<br>d703 |

| | | | | | |
|---|---|---|---|---|---|
| | | | | | d704<br>d705<br>d706<br>d709<br>d710<br>d714<br>d716<br>d717<br>d702<br>d708<br>d711<br>d719<br>d721<br>pa706<br>d720 |

**Table S2.** Original and recoded response options for religiosity items.

| Measure | Original response options | Recoded response options | Frequency Mothers | Frequency Partners |
|---|---|---|---|---|
| 1) Do you believe in God or in some divine power? | **Yes** | **Yes** | **49.8%** | **37.0%** |
| | **Not sure** | **Not sure** | **35.3%** | **34.4%** |
| | **No** | **No** | **14.9%** | **28.6%** |
| 2) Do you feel that God (or some divine power) has helped you at any time? | **Yes** | **Yes** | **33.9%** | **25.3%** |
| | **Not sure** | **Not sure** | **37.9%** | **32.3%** |
| | **No** | **No** | **28.3%** | **42.4%** |
| 3) Would you appeal to God for help if you were in trouble? | **Yes** | **Yes** | **46.6%** | **36.1%** |
| | **Not sure** | **Not sure** | **31.4%** | **27.5%** |
| | **No** | **No** | **22.1%** | **36.4%** |
| 4) How long have you had this particular faith? | **All my life[1]** | **Long-time (non)believer[1]** | **96.9%** | **96.7%** |
| | **More than 5 years[1]** | **Recent convert[2]** | | |
| | **3-5 years[2]** | | **3.1%** | **3.3%** |
| | **1-2 years[2]** | | | |
| | **Less than a year[2]** | | | |
| | **No response[1]** | | | |
| 5) Do you go to a place of worship? | **Yes, at least once a week[1]** | **Religious attendance[1]** | **14.3%** | **10.4%** |
| | **Yes, at least once a month[1]** | **Occasional attendance[2]** | | |
| | **Yes, at least once a year[2]** | | **85.7%** | **89.6%** |
| | **Not at all[2]** | | | |
| | **No response[2]** | | | |
| 6) Do you obtain help and support from leaders [other members] of your [other] religious group? | **Yes[1]** | **Yes[1]** | **11.3%** | **8.0%** |
| | **No[2]** | **No[2]** | **88.7%** | **92.0%** |
| | **No response[2]** | | | |

**Note.** Superscript indicates recoding pattern.

**Table S3.** Prevalence of psychological problems in the ALSPAC study population.

| Disorder derived from DAWBA | Derivation of dichotomous outcome variables from list of symptoms in DAWBA and examples of items in DAWBA relating to each outcome | Prevalence in ALSPAC study population |
|---|---|---|
| Separation anxiety | Any separation anxiety symptom(s) ‘a lot more than others’ compared with ‘no more than others’’ or ‘a little more than others’ for example, has he/she worried about sleeping alone? | 6.9% |
| Social phobia | Any social fears ‘‘a lot’’ compared with ‘none’ ’a little,’ or ‘hasn’t done this in the last month’ for example, has he/she been afraid of meeting new people? | 5.5% |
| Specific phobia | Any particular fears ‘a great deal’ compared with ‘‘quite a lot’, ‘only a little’ or ‘not at all’ for example, is he/she scared of the dark? | 12.4% |
| Generalized anxiety disorder | Any of the worries ‘often’ compared with ‘sometimes’ or ‘not at all’ for example, does he/she worry a lot about schoolwork, homework or tests/examinations? | 8.6% |
| Any depressive disorder | Any mood symptoms compared with none for example, did he/she think about death a lot? | 11.6% |
| Any ADHD disorder | Any attention/activity problems ‘a lot more than others’ compared with ‘a little more than others’ or ‘none’ for example, does he/she often fidget? Is he/she easily distracted? | 11.1% |
| Oppositional-defiant disorder | Any of the behaviours ‘a lot more than others’ compared with ‘no more than others’ or ‘a little more than others’ for example, has he/she had severe temper tantrums? | 6.2% |
| Obsessive compulsive disorder | Any of the behaviours ‘sometimes/often’ compared with ‘never’, for example have they engaged in excessive cleaning e.g., hand washing, baths, showers, toothbrushing etc? | 3.9% |
| Conduct disorder | Any of the behaviours ‘sometimes/often’ compared with ‘No, for example have they threatened someone, initiated a fight. | 6.7% |

**Table S4.** Prevalence of psychosocial problems in the ALSPAC study population.

| Psychosocial outcome variable | Derivation of dichotomous outcome variables | Prevalence in ALSPAC study population |
|---|---|---|
| **Antisocial behaviour** | Any antisocial behaviour was taken as an indicator. Examples include "Have you ever taken something from a shop without paying for it?" | 22.6% |
| **Unhappy with friends** | A score of 6 or more was coded as a high score. Examples include "Are you happy with the number of friends you've got?" | 17.1% |
| **Low scholastic competence** | Scores equal to or less than 14 were coded as low scores. Examples include "some children do very well at their classwork" | 29.7% |
| **Low self-worth** | Scores equal to or less than 14 were coded as low scores. Examples include "some children are happy with themselves as a person" | 9.3% |
| **Overt victim of bullying** | If child responded *yes* to any bullying event, he/she was classified as overt or relational bully/victim if bullying event happened frequently (several times a month) or very frequently (several times a week). For example, "has child hit been hit/beaten up?" | 34.2% |
| **Overt bully** | If child responded *yes* to any bullying event, he/she was classified as overt or relational bully/victim if bullying event happened frequently (several times a month) or very frequently (several times a week). For example, "has child hit/beaten up others?" | 7.0% |
| **Relational victim of bullying** | If child responded *yes* to any bullying event, he/she was classified as overt or relational bully/victim if bullying event happened frequently (several times a month) or very frequently (several times a week). For example, "has child had games spoilt?" | 16.4% |
| **Relational bully** | If child responded *yes* to any bullying event, he/she was classified as overt or relational bully/victim if bullying event happened frequently (several times a month) or very frequently (several times a week). For example, "has child spoilt other children's games?" | 2.4% |

**Table S5.** Weighted odds ratios and 95% confidence intervals for the associations between maternal religious latent class and offspring mental health at age 7, with the Agnostic class as the reference class.

| | OR (CI) Model 1 | OR (CI) Model 2 | OR (CI) Model 3 | OR (CI) Model 4 | OR (CI) Model 5 |
|---|---|---|---|---|---|
| **ADHD** | | | | | |
| Agnostic | 1.00 ref | 1.00 ref | 1.00 ref | 1.00 ref | 1.00 ref |
| Highly religious | 1.29(1.00,1.58) | 1.30(1.01,1.59) | 1.38(1.08,1.67) | 1.35(1.06,1.65) | 1.34(1.05,1.64) |
| Moderately religious | 1.16(0.90,1.41) | 1.16(0.91,1.42) | 1.19(0.94,1.45) | 1.19(0.93,1.45) | 1.17(0.91,1.43) |
| Atheist | 1.44(1.18,1.71) | 1.43(1.16,1.69) | 1.42(1.16,1.69) | 1.40(1.13,1.67) | 1.41(1.14,1.68) |
| P - value | .043 | .052 | .040 | .057 | .053 |
| **Conduct disorder** | | | | | |
| Agnostic | 1.00 ref | 1.00 ref | 1.00 ref | 1.00 ref | 1.00 ref |
| Highly religious | 1.35(0.98,1.72) | 1.36(0.99,1.73) | 1.40(1.03,1.78) | 1.38(1.00,1.76) | 1.37(0.99,1.75) |
| Moderately religious | 1.19(0.86,1.53) | 1.21(0.88,1.54) | 1.24(0.91,1.58) | 1.23(0.89,1.56) | 1.22(0.89,1.56) |
| Atheist | 1.55(1.22,1.89) | 1.51(1.18,1.85) | 1.48(1.15,1.82) | 1.45(1.12,1.79) | 1.46(1.12,1.80) |
| P - value | .067 | .090 | .102 | .134 | .133 |
| **Depression** | | | | | |
| Agnostic | 1.00 ref | 1.00 ref | 1.00 ref | 1.00 ref | 1.00 ref |
| Highly religious | 1.51(1.24,1.77) | 1.51(1.24,1.77) | 1.43(1.16,1.71) | 1.41(1.13,1.68) | 1.40(1.13,1.68) |
| Moderately religious | 1.50(1.26,1.73) | 1.50(1.27,1.74) | 1.50(1.27,1.74) | 1.49(1.26,1.73) | 1.48(1.24,1.71) |
| Atheist | 1.17(0.90,1.44) | 1.16(0.90,1.43) | 1.15(0.88,1.42) | 1.13(0.86,1.40) | 1.13(0.86,1.40) |
| P - value | .001 | .001 | .002 | .003 | .004 |

| | | | | | |
|---|---|---|---|---|---|
| **General anxiety** | | | | | |
| Agnostic | 1.00 ref | 1.00 ref | 1.00 ref | 1.00 ref | 1.00 ref |
| Highly religious | 1.21(0.89,1.53) | 1.21(0.89,1.53) | 1.23(0.91,1.56) | 1.23(0.90,1.55) | 1.22(0.90,1.55) |
| Moderately religious | 1.44(1.18,1.70) | 1.44(1.18,1.71) | 1.46(1.20,1.73) | 1.45(1.19,1.72) | 1.43(1.17,1.70) |
| Atheist | 1.24(0.94,1.53) | 1.24(0.94,1.53) | 1.24(0.94,1.54) | 1.23(0.93,1.53) | 1.24(0.94,1.53) |
| P - value | .056 | .055 | .045 | .050 | .066 |
| **OCD** | | | | | |
| Agnostic | 1.00 ref | 1.00 ref | 1.00 ref | 1.00 ref | 1.00 ref |
| Highly religious | 1.60(1.21,2.00) | 1.60(1.21,1.99) | 1.58(1.19,1.97) | 1.55(1.16,1.95) | 1.53(1.13,1.92) |
| Moderately religious | 0.65(0.19,1.11) | 0.65(0.19,1.11) | 0.65(0.20,1.11) | 0.65(0.19,1.11) | 0.64(0.18,1.10) |
| Atheist | 1.04(0.63,1.46) | 1.05(0.63,1.47) | 1.04(0.62,1.47) | 1.03(0.61,1.45) | 1.03(0.62,1.45) |
| P - value | .004 | .004 | .006 | .007 | .008 |
| **Social phobia** | | | | | |
| Agnostic | 1.00 ref | 1.00 ref | 1.00 ref | 1.00 ref | 1.00 ref |
| Highly religious | 1.39(1.01,1.76) | 1.39(1.01,1.76) | 1.37(0.99,1.75) | 1.34(0.96,1.72) | 1.33(0.95,1.71) |
| Moderately religious | 1.25(0.91,1.58) | 1.25(0.91,1.58) | 1.26(0.93,1.60) | 1.25(0.91,1.58) | 1.24(0.90,1.57) |
| Atheist | 1.04(0.65,1.43) | 1.04(0.65,1.43) | 1.02(0.63,1.41) | 1.00(0.61,1.39) | 1.01(0.62,1.40) |
| P - value | .255 | .254 | .259 | .287 | .328 |
| **Specific phobia** | | | | | |
| Agnostic | 1.00 ref | 1.00 ref | 1.00 ref | 1.00 ref | 1.00 ref |
| Highly religious | 1.01(0.74,1.28) | 1.01(0.74,1.28) | 1.06(0.79,1.33) | 1.06(0.78,1.33) | 1.06(0.78,1.33) |

| | | | | | |
|---|---|---|---|---|---|
| Moderately religious | 1.03(0.80,1.26) | 1.03(0.81,1.26) | 1.05(0.82,1.28) | 1.05(0.82,1.28) | 1.05(0.82,1.28) |
| Atheist | 0.86(0.60,1.12) | 0.85(0.59,1.11) | 0.85(0.58,1.11) | 0.84(0.58,1.11) | 0.84(0.58,1.11) |
| P - value | .585 | .499 | .374 | .368 | .383 |
| **ODD** | | | | | |
| Agnostic | 1.00 ref | 1.00 ref | 1.00 ref | 1.00 ref | 1.00 ref |
| Highly religious | 1.73(1.37,2.10) | 1.74(1.38,2.10) | 1.78(1.41,2.14) | 1.74(1.37,2.10) | 1.72(1.35,2.08) |
| Moderately religious | 1.39(1.06,1.72) | 1.40(1.06,1.73) | 1.43(1.10,1.77) | 1.41(1.07,1.75) | 1.38(1.04,1.72) |
| Atheist | 1.44(1.08,1.79) | 1.42(1.07,1.78) | 1.40(1.04,1.76) | 1.37(1.01,1.73) | 1.39(1.02,1.75) |
| P - value | .020 | .020 | .015 | .022 | .027 |
| **Separation anxiety** | | | | | |
| Agnostic | 1.00 ref | 1.00 ref | 1.00 ref | 1.00 ref | 1.00 ref |
| Highly religious | 1.19(0.83,1.55) | 1.19(0.83,1.55) | 1.19(0.82,1.56) | 1.18(0.81,1.55) | 1.18(0.81,1.56) |
| Moderately religious | 1.23(0.92,1.54) | 1.24(0.93,1.55) | 1.26(0.95,1.57) | 1.25(0.94,1.57) | 1.25(0.94,1.56) |
| Atheist | 1.09(0.75,1.44) | 1.08(0.74,1.43) | 1.06(0.71,1.41) | 1.05(0.70,1.40) | 1.05(0.70,1.40) |
| P - value | .552 | .530 | .469 | .479 | .775 |

**Note.** *Model 2* adjusts for maternal age, *Model 3* for maternal age and SEP, *Model 4* for maternal age, SEP and ACE, and *Model 5* for maternal age, SEP, ACE, and maternal mental health. P values are omnibus p-values based on a Wald test with 3 d.f.

**Table S6.** Weighted odds ratios and 95% confidence intervals for the associations between maternal religious latent class and offspring psychosocial outcomes at age 8, with the Agnostic class as the reference class.

| | **OR (CI)** **Model 1** | **OR (CI)** **Model 2** | **OR (CI)** **Model 3** | **OR (CI)** **Model 4** | **OR (CI)** **Model 5** |
|---|---|---|---|---|---|
| **Antisocial behaviour** | | | | | |
| Agnostic | 1.00 ref | 1.00 ref | 1.00 ref | 1.00 ref | 1.00 ref |
| Highly religious | 1.29(1.06,1.52) | 1.29(1.06,1.52) | 1.31(1.08,1.54) | 1.29(1.05,1.52) | 1.29(1.06,1.52) |
| Moderately religious | 1.24(1.04,1.44) | 1.24(1.04,1.44) | 1.24(1.04,1.44) | 1.23(1.03,1.43) | 1.24(1.04,1.44) |
| Atheist | 1.42(1.21,1.64) | 1.42(1.20,1.63) | 1.41(1.19,1.63) | 1.40(1.18,1.61) | 1.40(1.18,1.61) |
| P - value | .008 | .009 | .009 | .014 | .013 |
| **Low scholastic competence** | | | | | |
| Agnostic | 1.00 ref | 1.00 ref | 1.00 ref | 1.00 ref | 1.00 ref |
| Highly religious | 0.96(0.74,1.17) | 0.96(0.74,1.18) | 1.04(0.82,1.26) | 1.03(0.81,1.25) | 1.04(0.82,1.26) |
| Moderately religious | 1.17(1.00,1.35) | 1.18(1.00,1.36) | 1.21(1.03,1.39) | 1.21(1.03,1.39) | 1.22(1.04,1.40) |
| Atheist | 1.10(0.90,1.30) | 1.10(0.90,1.30) | 1.12(0.92,1.32) | 1.12(0.91,1.32) | 1.12(0.92,1.32) |
| P - value | .241 | .225 | .223 | .215 | .184 |
| **Low self-worth** | | | | | |
| Agnostic | 1.00 ref | 1.00 ref | 1.00 ref | 1.00 ref | 1.00 ref |
| Highly religious | 1.26(0.99,1.52) | 1.26(0.99,1.52) | 1.25(0.98,1.52) | 1.26(0.99,1.53) | 1.27(1.00,1.54) |
| Moderately religious | 1.15(0.91,1.38) | 1.16(0.92,1.39) | 1.15(0.92,1.39) | 1.15(0.92,1.39) | 1.17(0.94,1.41) |
| Atheist | 1.17(0.91,1.43) | 1.16(0.91,1.42) | 1.15(0.89,1.41) | 1.15(0.90,1.41) | 1.16(0.90,1.42) |
| P - value | .324 | .321 | .356 | .339 | .289 |

| | | | | | |
|---|---|---|---|---|---|
| **Overt victim** | | | | | |
| Agnostic | 1.00 ref | 1.00 ref | 1.00 ref | 1.00 ref | 1.00 ref |
| Highly religious | 0.91(0.71,1.11) | 0.91(0.71,1.11) | 0.88(0.68,1.09) | 0.89(0.69,1.10) | 0.89(0.69,1.10) |
| Moderately religious | 0.95(0.78,1.12) | 0.95(0.78,1.12) | 0.94(0.77,1.11) | 0.94(0.77,1.12) | 0.95(0.78,1.12) |
| Atheist | 0.94(0.75,1.14) | 0.95(0.76,1.14) | 0.94(0.75,1.14) | 0.95(0.76,1.15) | 0.96(0.76,1.15) |
| P - value | .827 | .826 | .669 | .733 | .743 |
| **Overt bully** | | | | | |
| Agnostic | 1.00 ref | 1.00 ref | 1.00 ref | 1.00 ref | 1.00 ref |
| Highly religious | 0.60(0.22,0.98) | 0.62(0.26,0.98) | 0.59(0.20,0.98) | 0.59(0.20,0.97) | 0.59(0.21,0.98) |
| Moderately religious | 0.74(0.38,1.09) | 0.74(0.40,1.09) | 0.72(0.37,1.08) | 0.72(0.37,1.08) | 0.73(0.38,1.09) |
| Atheist | 0.57(0.20,0.94) | 0.58(0.19,0.97) | 0.58(0.21,0.95) | 0.58(0.21,0.95) | 0.58(0.21,0.95) |
| P - value | .013 | .014 | .013 | .012 | .013 |
| **Relational bully** | | | | | |
| Agnostic | 1.00 ref | 1.00 ref | 1.00 ref | 1.00 ref | 1.00 ref |
| Highly religious | 0.71(0.13,1.29) | 0.71(0.13,1.29) | 0.67(0.09,1.25) | 0.64(0.06,1.23) | 0.65(0.07,1.24) |
| Moderately religious | 1.03(0.48,1.59) | 1.02(0.47,1.57) | 1.01(0.46,1.56) | 1.01(0.45,1.57) | 1.05(0.48,1.61) |
| Atheist | 0.90(0.30,1.49) | 0.91(0.31,1.51) | 0.90(0.31,1.49) | 0.89(0.30,1.49) | 0.90(0.31,1.49) |
| P - value | .642 | .649 | .535 | .468 | .458 |
| **Relational victim** | | | | | |
| Agnostic | 1.00 ref | 1.00 ref | 1.00 ref | 1.00 ref | 1.00 ref |
| Highly religious | 0.77(0.52,1.02) | 0.77(0.52,1.01) | 0.72(0.47,0.97) | 0.74(0.49,0.99) | 0.74(0.49,0.99) |
| Moderately religious | 0.94(0.72,1.16) | 0.93(0.71,1.15) | 0.91(0.69,1.14) | 0.92(0.70,1.14) | 0.92(0.70,1.15) |

| | | | | | |
|---|---|---|---|---|---|
| Atheist | 1.08(0.82,1.33) | 1.09(0.83,1.34) | 1.08(0.82,1.33) | 1.09(0.83,1.34) | 1.09(0.84,1.35) |
| P - value | .103 | .087 | .033 | .045 | .045 |
| **Unhappy with friends** | | | | | |
| Agnostic | 1.00 ref | 1.00 ref | 1.00 ref | 1.00 ref | 1.00 ref |
| Highly religious | 1.15(0.91,1.39) | 1.15(0.91,1.39) | 1.17(0.93,1.42) | 1.16(0.91,1.40) | 1.15(0.91,1.40) |
| Moderately religious | 0.93(0.72,1.14) | 0.93(0.72,1.14) | 0.94(0.73,1.16) | 0.94(0.73,1.16) | 0.94(0.72,1.15) |
| Atheist | 0.77(0.51,1.02) | 0.77(0.52,1.02) | 0.77(0.52,1.02) | 0.76(0.51,1.01) | 0.76(0.51,1.01) |
| P - value | .047 | .048 | .040 | .042 | .043 |

**Note.** *Model 2* adjusts for maternal age, *Model 3* for maternal age and SEP, *Model 4* for maternal age, SEP, and ACE, and *Model 5* for maternal age, SEP, ACE, and maternal mental health. P values are omnibus p-values based on a Wald test with 3 d.f.

**Table S7.** Unweighted odds ratios and 95% confidence intervals for the associations between parental religious latent class and offspring mental health, with the Agnostic class as the reference class.

| | **OR (CI) Model 1** | **OR (CI) Model 2** | **OR (CI) Model 3** | **OR (CI) Model 4** | **OR (CI) Model 5** |
|---|---|---|---|---|---|
| ADHD | | | | | |
| Agnostic | 1.00 ref | 1.00 ref | 1.00 ref | 1.00 ref | 1.00 ref |
| Highly religious | 1.22(0.93,1.50) | 1.25(0.96,1.54) | 1.34(1.05,1.62) | 1.31(1.02,1.59) | 1.30(1.01,1.59) |
| Moderately religious | 1.21(0.96,1.45) | 1.22(0.98,1.46) | 1.24(1.00,1.49) | 1.23(0.99,1.48) | 1.22(0.98,1.47) |
| Atheist | 1.44(1.19,1.69) | 1.43(1.18,1.69) | 1.40(1.15,1.65) | 1.37(1.12,1.63) | 1.38(1.13,1.64) |
| P - value | .040 | .040 | .044 | .069 | .068 |
| Conduct disorder | | | | | |
| Agnostic | 1.00 ref | 1.00 ref | 1.00 ref | 1.00 ref | 1.00 ref |
| Highly religious | 1.30(0.95,1.66) | 1.38(1.03,1.74) | 1.45(1.09,1.82) | 1.41(1.04,1.77) | 1.40(1.04,1.77) |
| Moderately religious | 1.17(0.85,1.48) | 1.20(0.89,1.51) | 1.22(0.91,1.54) | 1.20(0.88,1.52) | 1.20(0.88,1.51) |
| Atheist | 1.54(1.23,1.86) | 1.52(1.21,1.83) | 1.44(1.13,1.76) | 1.40(1.08,1.72) | 1.40(1.09,1.72) |
| P - value | .049 | .051 | .077 | .124 | .124 |
| Depression | | | | | |
| Agnostic | 1.00 ref | 1.00 ref | 1.00 ref | 1.00 ref | 1.00 ref |
| Highly religious | 1.48(1.22,1.75) | 1.57(1.30,1.84) | 1.47(1.20,1.74) | 1.44(1.16,1.71) | 1.44(1.17,1.71) |
| Moderately religious | 1.55(1.32,1.78) | 1.59(1.36,1.82) | 1.59(1.36,1.82) | 1.57(1.34,1.80) | 1.56(1.33,1.79) |
| Atheist | 1.21(0.95,1.47) | 1.20(0.94,1.46) | 1.18(0.92,1.44) | 1.15(0.89,1.41) | 1.15(0.89,1.41) |
| P - value | .001 | .000 | .000 | .000 | .001 |
| GAD | | | | | |
| Agnostic | 1.00 ref | 1.00 ref | 1.00 ref | 1.00 ref | 1.00 ref |
| Highly religious | 1.15(0.84,1.47) | 1.21(0.90,1.52) | 1.21(0.89,1.53) | 1.19(0.88,1.51) | 1.19(0.87,1.51) |
| Moderately religious | 1.43(1.18,1.69) | 1.46(1.21, 1.71) | 1.47(1.22,1.73) | 1.46(1.21,1.71) | 1.44(1.19,1.70) |
| Atheist | 1.24(0.97,1.52) | 1.23(0.95,1.51) | 1.21(0.93,1.49) | 1.20(0.91,1.48) | 1.20(0.92,1.48) |
| P - value | 0.047 | 0.032 | 0.028 | 0.034 | 0.045 |
| OCD | | | | | |
| Agnostic | 1.00 ref | 1.00 ref | 1.00 ref | 1.00 ref | 1.00 ref |
| Highly religious | 1.53(1.15,1.92) | 1.71(1.33,2.10) | 1.58(1.19,1.96) | 1.55(1.16,1.93) | 1.52(1.14,1.90) |
| Moderately religious | 0.63(0.19,1.08) | 0.67(0.22,1.11) | 0.66(0.22,1.10) | 0.65(0.20,1.10) | 0.65(0.20,1.09) |
| Atheist | 1.10(0.70,1.50) | 1.07(0.67,1.47) | 1.07(0.67,1.47) | 1.05(0.64,1.45) | 1.06(0.65,1.46) |
| P - value | .005 | .002 | .006 | .007 | .007 |

| | | | | | |
|---|---|---|---|---|---|
| Social phobia | | | | | |
| Agnostic | 1.00 ref | 1.00 ref | 1.00 ref | 1.00 ref | 1.00 ref |
| Highly religious | 1.32(0.95,1.68) | 1.31(0.95,1.68) | 1.32(0.94,1.69) | 1.27(0.89,1.65) | 1.26(0.88,1.64) |
| Moderately religious | 1.25(0.93,1.57) | 1.25(0.93,1.57) | 1.26(0.94,1.58) | 1.24(0.92,1.56) | 1.23(0.91,1.55) |
| Atheist | 1.00(0.64,1.36) | 1.00(0.63,1.37) | 0.97(0.60,1.34) | 0.94(0.56,1.31) | 0.94(0.57,1.31) |
| P - value | .272 | .287 | .225 | .258 | .304 |
| Specific phobia | | | | | |
| Agnostic | 1.00 ref | 1.00 ref | 1.00 ref | 1.00 ref | 1.00 ref |
| Highly religious | 0.99(0.72,1.26) | 1.05(0.78,1.32) | 1.10(0.82,1.37) | 1.09(0.81,1.36) | 1.09(0.81,1.36) |
| Moderately religious | 1.07(0.86,1.29) | 1.10(0.88,1.32) | 1.11(0.89,1.33) | 1.11(0.89,1.33) | 1.11(0.89,1.33) |
| Atheist | 0.95(0.71,1.20) | 0.94(0.70,1.19) | 0.93(0.68,1.17) | 0.92(0.68,1.17) | 0.92(0.68,1.17) |
| P - value | .813 | .639 | .468 | .465 | .492 |
| ODD | | | | | |
| Agnostic | 1.00 ref | 1.00 ref | 1.00 ref | 1.00 ref | 1.00 ref |
| Highly religious | 1.60(1.25,1.95) | 1.66(1.31,2.01) | 1.73(1.38,2.09) | 1.68(1.33,2.04) | 1.66(1.3,2.02) |
| Moderately religious | 1.39(1.07,1.71) | 1.41(1.10,1.73) | 1.44(1.12,1.76) | 1.41(1.09,1.73) | 1.39(1.07,1.71) |
| Atheist | 1.47(1.13,1.80) | 1.41(1.08,1.75) | 1.38(1.05,1.72) | 1.35(1.01,1.68) | 1.36(1.02,1.69) |
| P - value | .031 | .020 | .014 | .024 | .030 |
| Separation anxiety | | | | | |
| Agnostic | | | | | |
| Highly religious | 1.18(0.82,1.54) | 1.16(0.81,1.52) | 1.16(0.81,1.52) | 1.19(0.82,1.56) | 1.18(0.81,1.55) |
| Moderately religious | 1.30(1.01,1.60) | 1.29(1.00,1.59) | 1.29(1.00,1.59) | 1.31(1.02,1.61) | 1.31(1.01,1.61) |
| Atheist | 1.19(0.87,1.52) | 1.20(0.87,1.52) | 1.20(0.87,1.52) | 1.15(0.82,1.48) | 1.14(0.81,1.47) |
| P - value | 0.361 | 0.387 | 0.343 | 0.357 | 0.359 |

**Table S8.** Unweighted odds ratios and 95% confidence intervals for the associations between parental religious latent class and offspring psychosocial outcomes, with the Agnostic class as the reference class.

| | OR (CI) Model 1 | OR (CI) Model 2 | OR (CI) Model 3 | OR (CI) Model 4 | OR (CI) Model 5 |
|---|---|---|---|---|---|
| Antisocial behaviour | | | | | |
| Agnostic | 1.00 ref | 1.00 ref | 1.00 ref | 1.00 ref | 1.00 ref |
| Highly religious | 1.28(1.05,1.50) | 1.27(1.05,1.50) | 1.30(1.07,1.52) | 1.28(1.05,1.50) | 1.28(1.05,1.50) |
| Moderately religious | 1.19(0.99,1.38) | 1.18(0.99,1.38) | 1.18(0.99,1.37) | 1.17(0.98,1.37) | 1.17(0.97,1.36) |
| Atheist | 1.38(1.17,1.58) | 1.38(1.17,1.58) | 1.35(1.14,1.56) | 1.34(1.14,1.55) | 1.35(1.14,1.55) |
| P - value | .014 | .015 | .019 | .026 | .025 |
| Low scholastic competence | | | | | |
| Agnostic | 1.00 ref | 1.00 ref | 1.00 ref | 1.00 ref | 1.00 ref |
| Highly religious | 0.97(0.76,1.19) | 1.00(0.78,1.21) | 1.07(0.86,1.29) | 1.07(0.85,1.28) | 1.07(0.85,1.28) |
| Moderately religious | 1.13(0.96,1.30) | 1.15(0.97,1.32) | 1.16(0.99,1.34) | 1.16(0.99,1.34) | 1.16(0.98,1.33) |
| Atheist | 1.15(0.96,1.34) | 1.15(0.96,1.35) | 1.15(0.96,1.35) | 1.15(0.96,1.35) | 1.16(0.97,1.36) |
| P - value | .318 | .305 | .324 | .319 | .315 |
| Low self-worth | | | | | |
| Agnostic | 1.00 ref | 1.00 ref | 1.00 ref | 1.00 ref | 1.00 ref |
| Highly religious | 1.31(1.05,1.56) | 1.32(1.06,1.58) | 1.32(1.06,1.59) | 1.33(1.07,1.60) | 1.33(1.06,1.59) |
| Moderately religious | 1.07(0.84,1.30) | 1.08(0.85,1.31) | 1.08(0.85,1.31) | 1.08(0.85,1.31) | 1.07(0.84,1.31) |
| Atheist | 1.22(0.97,1.47) | 1.22(0.98,1.47) | 1.20(0.95,1.44) | 1.20(0.95,1.45) | 1.21(0.96,1.46) |
| P - value | .153 | .141 | .166 | .160 | .152 |
| Overt victim | | | | | |
| Agnostic | 1.00 ref | 1.00 ref | 1.00 ref | 1.00 ref | 1.00 ref |
| Highly religious | 0.94(0.74,1.14) | 0.91(0.71,1.11) | 0.88(0.68,1.09) | 0.90(0.70,1.10) | 0.90(0.70,1.10) |
| Moderately religious | 0.97(0.80,1.14) | 0.95(0.78,1.12) | 0.94(0.78,1.11) | 0.95(0.78,1.12) | 0.95(0.79,1.12) |
| Atheist | 0.93(0.75,1.12) | 0.93(0.74,1.11) | 0.94(0.75,1.12) | 0.94(0.76,1.13) | 0.94(0.75,1.12) |
| P - value | .878 | .767 | .657 | .743 | .748 |
| Overt bully | | | | | |
| Agnostic | 1.00 ref | 1.00 ref | 1.00 ref | 1.00 ref | 1.00 ref |
| Highly religious | 0.59(0.22,0.96) | 0.58(0.21,0.96) | 0.56(0.18,0.94) | 0.56(0.18,0.93) | 0.56(0.18,0.94) |
| Moderately religious | 0.73(0.39,1.07) | 0.73(0.38,1.07) | 0.72(0.38,1.06) | 0.72(0.38,1.06) | 0.72(0.38,1.07) |
| Atheist | 0.58(0.22,0.93) | 0.58(0.22,0.93) | 0.59(0.23,0.94) | 0.58(0.23,0.94) | 0.58(0.23,0.94) |

| | | | | | |
|---|---|---|---|---|---|
| P - value | .007 | .006 | .006 | .005 | .005 |
| Relational bully | | | | | |
| Agnostic | 1.00 ref | 1.00 ref | 1.00 ref | 1.00 ref | 1.00 ref |
| Highly religious | 0.77(0.20,1.35) | 0.76(0.17,1.34) | 0.71(0.13,1.29) | 0.69(0.10,1.28) | 0.69(0.10,1.28) |
| Moderately religious | 0.94(0.41,1.47) | 0.93(0.40,1.45) | 0.92(0.39,1.45) | 0.93(0.40,1.46) | 0.96(0.43,1.49) |
| Atheist | 0.89(0.32,1.46) | 0.88(0.31,1.45) | 0.90(0.34,1.46) | 0.89(0.33,1.45) | 0.88(0.32,1.44) |
| P - value | .855 | .828 | .724 | .679 | .664 |
| Relational victim | | | | | |
| Agnostic | 1.00 ref | 1.00 ref | 1.00 ref | 1.00 ref | 1.00 ref |
| Highly religious | 0.83(0.59,1.08) | 0.80(0.56,1.05) | 0.76(0.51,1.01) | 0.78(0.53,1.03) | 0.78(0.53,1.03) |
| Moderately religious | 0.95(0.74,1.17) | 0.94(0.72,1.15) | 0.92(0.71,1.13) | 0.92(0.71,1.14) | 0.93(0.72,1.15) |
| Atheist | 1.05(0.80,1.29) | 1.04(0.80,1.28) | 1.04(0.80,1.28) | 1.05(0.81,1.29) | 1.04(0.80,1.28) |
| P - value | .376 | .255 | .109 | .149 | .173 |
| Unhappy with friends | | | | | |
| Agnostic | 1.00 ref | 1.00 ref | 1.00 ref | 1.00 ref | 1.00 ref |
| Highly religious | 1.12(0.88,1.35) | 1.11(0.87,1.35) | 1.13(0.89,1.38) | 1.12(0.88,1.36) | 1.12(0.88,1.36) |
| Moderately religious | 0.92(0.71,1.12) | 0.91(0.70,1.12) | 0.92(0.72,1.13) | 0.92(0.71,1.13) | 0.92(0.71,1.13) |
| Atheist | 0.80(0.56,1.04) | 0.80(0.56,1.04) | 0.79(0.55,1.03) | 0.78(0.55,1.02) | 0.79(0.55,1.02) |
| P - value | .113 | .120 | .082 | .088 | .092 |
